# Effect of temperature and precipitation on the daily new cases and daily new death in seven cities around the globe

**DOI:** 10.1101/2020.10.03.20206227

**Authors:** Amar Prashad Chaudhary, Adna Nelson K, Harish S, Mydhily S, Chaitanya KJ, Lovely Nayak, Chiranjibi Sah

**Affiliations:** Rajiv Gandhi University of Health Sciences

**Author notes:** Corresponding Author: Amar Prashad Chaudhary, Email ID.

**Keywords:** COVID-19, SARS-COV-2, Temperature, Precipitation, Transmission, death

## Abstract

**Background:** This study was done to understand the effect of temperature and precipitation in COVID-19.

**Objective:** To study the effect of temperature and precipitation on transmission of COVID-19.

To study the effect of temperature and precipitation on daily death of COVID-19.

**Methodology:** We collected 3 consecutive month data of seven cities around the world which were effected most by the COVID-19. Data included weather variables i.e temperature (average temperature, maximum temperature and minimum temperature), precipitation, daily new cases and daily new death.

**Conclusion:** Increase in average temperature reduces daily death and increase in maximum temperature reduces transmission.

## INTRODUCTION

Covid19 is the third epidemic caused by coronavirus that the world has come across now. The first epidemic was during 2002–2003 in Guangdong province of China. The disease was termed ‘severe acute respiratory syndrome’ (SARS) and was caused by a then newly identified coronavirus that was named as SARS coronavirus. The second epidemic was in 2012 in the Middle East and the disease was termed ‘Middle East respiratory syndrome’ and was caused by a new virus called MERS. It was by the end of 2019 that the third epidemic originated in Wuhan, Hubei province in China and is caused by a new type of coronavirus that the World Health Organization named SARS-COV-2.(Felipe *et al*., 2020) The resultant disease was named ‘coronavirus disease’ (COVID-19). World Health Organization confirmed COVID-19 to be a pandemic on March 11, 2020 when they reported 118,319 confirmed cases and 4292 deaths all over the world by March 11,2020.(Ma *et al*., 2020)

The virus infects humans and is believed to infect animals, mainly pigs and bats and is confirmed to have human to human transmissibility.(Adnan *et al*., 2020; Felipe *et al*., 2020) The clinical manifestation of the infection mainly includes respiratory insufficiencies or syndrome with the severity ranging from mild to lethal and sometimes is asymptomatic. Most common symptoms include fever, dry cough and tiredness. Other observed symptoms include body aches, nausea, vomiting, sore throat, loss of taste, difficulty in breathing etc.

South East Asia was expected to be the region that would be most affected due to its geographical proximity to Wuhan but Tehran, Milan, Madrid, New York and London were the regions that were affected the most.(Sethwala *et al*., 2020) This led to questions about the effect of environmental factors such as temperature, humidity, precipitation and ultraviolet radiation on the viability and rate of transmission of SARS-COV-2.

Research has reported that a seasonal cyclic repetition was commonly observed in viral respiratory diseases. It was observed that viral respiratory infections are predominant during winter or colder seasons.(Weston and Frieman, 2019) For example, influenza outbreaks occur every winter in temperate regions. Also, a study in China that was based on SARS in Hong Kong, Guangzhou, Beijing, and Taiyuan indicated that the outbreaks were associated with variations in temperature.

It was confirmed that the rate of viral transmission was positively associated with temperature and humidity. It was also observed that the variations in temperature also had undeniable impact on the mortality rates due to the virus.(Wu *et al*., 2020)

Experiments on coronaviruses in controlled environments showed that conditions that encourage the survival of the virus are temperatures around 4°C and relative humidity between 20-80%. It was found that in these conditions, coronaviruses can survive up to 28 days. It was also found that inactivation of the viruses occurred rapidly above 20°C. When temperatures reach close to 40°C, the coronaviruses last only a few hours.(Casanova *et al*., 2010) It was suggested that the coronaviruses transmit more easily during cold weathers.(Kifer *et al*., 2020)

Even in the case of SARS-COV-2, meteorological factors seem to influence the spread of the virus.(Xu *et al*., 2020) It was observed that the highest outbreaks were in colder regions when compared to warmer regions.(Felipe *et al*., 2020; Ma *et al*., 2020) It was also advised that drinking warm water frequently, hot water gargle and avoiding cold food items can decrease chances of getting affected by the corona virus. This further suggests the possible effects of temperature on the viability of the virus.

A published laboratory study by Chin et al (2020) reported that SARS-COV-2 was highly stable at 4 °C but sensitive to heat. The survival time of the virus was shortened to 5 mins as the incubation temperature increased to 70 °C.(H Chin *et al*., 2020)

Assuming that different climatic conditions affect transmission of SARS-COV-2 differently, it is important to find the association between minimum temperature, maximum temperature, average temperature, precipitation and COVID-19 incident cases and COVID-19 deaths.

## Methodology

### Selection of city

The cities were selected on the basis of the temperature range and outburst of the COVID-19 in big cities around various part of globe to study the effect of temperature in wide range.

### Data collection

Metrological data i.e average daily temperature, minimum temperature, maximum temperature, precipitation and the COVID-19 daily new case and daily death of seven different cities belonging to six different countries was taken. The data was collected of three consecutive month during the period of COVID-19 outburst in those cities. All the details of the cities along with its country, time period of data collected and the public domain from where the data was collected is given below in detail in table 1.

**Table 1.**
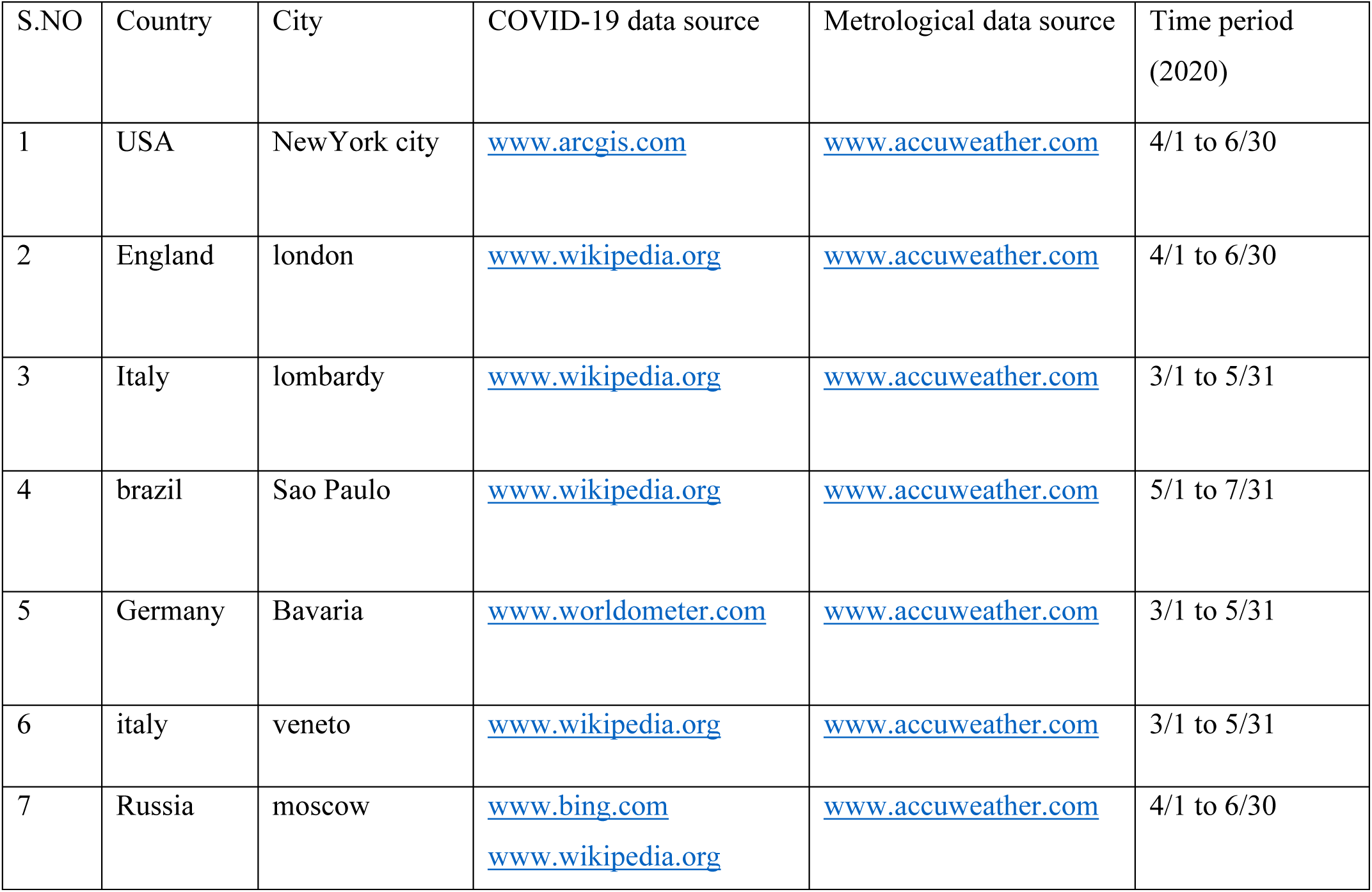

### Data processing strategy

For this study, two outcome research was considered. First outcome was effect of metrological parameters on transmission and second was effect of meteorological parameters on daily deaths. Two estimates were tested for outcomes. First estimates included the average temperature and precipitation while second estimate included minimum temperature, maximum temperature, average temperature and precipitation.

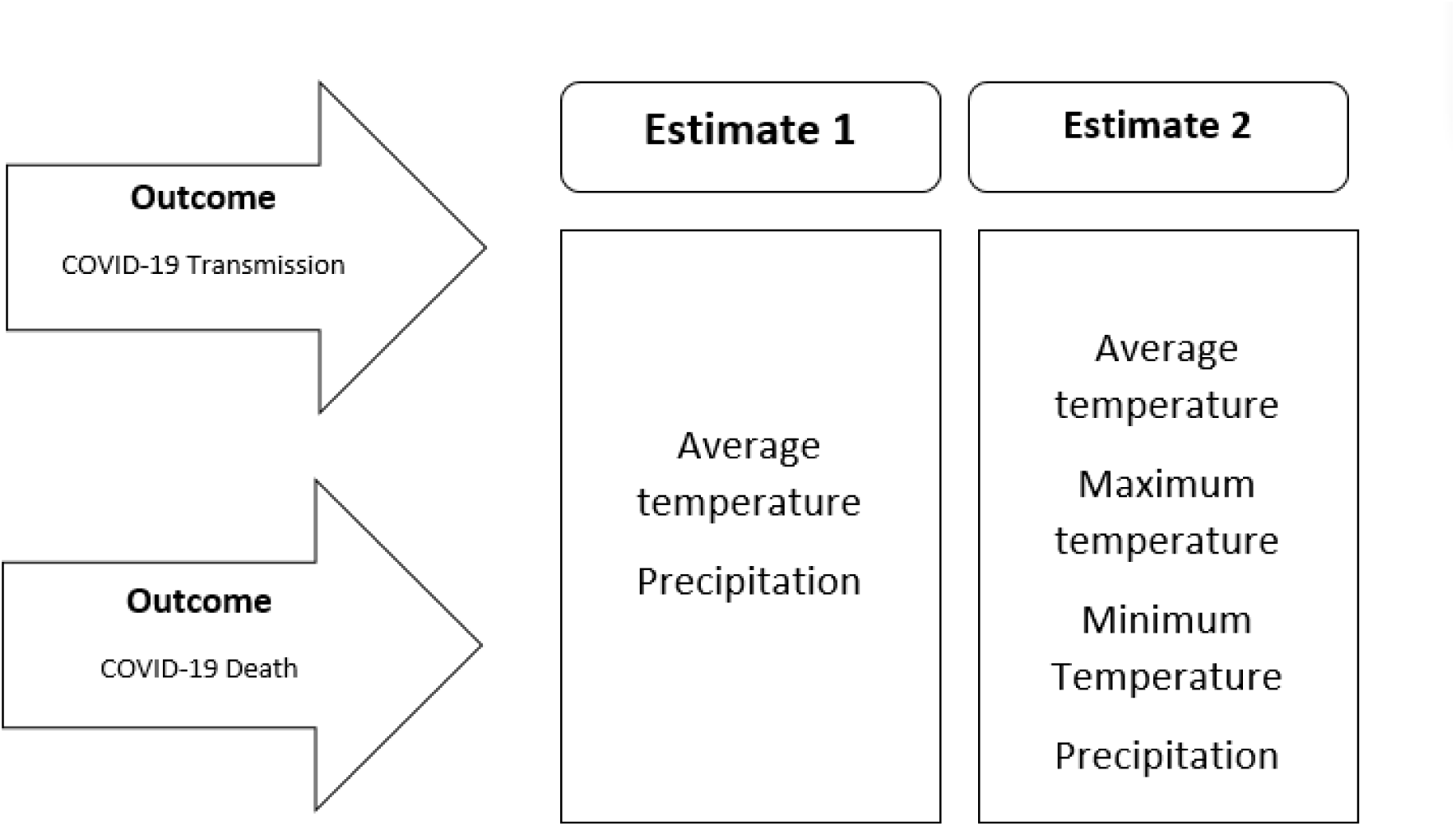

### Model

To estimate the association of daily new cases and daily deaths with metrological parameters like temperature (high temperature, low temperature, average temperature) and precipitation, we used common panel data regression model. The equation developed by panel data model using the give data is

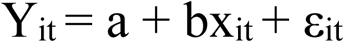

Where **Y**_***it***_ is the dependent variable (total number of infections or total number of deaths) for 6 countries **‘*i*’** over the period **“*t*”** in 2020; *a* is a constant; and **X**_**it**_ is a vector that includes all explanatory variables of the model, including the variable of interest, that is, the mean temperature and **ε**_**it**,_ which is the error term of this equation that includes all factors that are associated with **Y**_***it***_ and are not included in the equation.

## Results

Table 2 shows the result of correlation analysis between the daily death and metrological parameter. The first estimated model shows negative correlation between daily death and metrological parameters i.e average temperature and precipitation. The p value helps to understand there remains the significant association between average daily temperature and daily death. With the inclusion of two other variable i.e maximum temperature and minimum temperature in second estimated model, there still remains the negative correlation between daily death and metrological parameter i.e average temperature and precipitation but significance of the association wasn’t seen. However, as per the results of first estimated model helps to understand that drop in the temperature increase the death among COVID-19 patients. This result helps to predict in summer season there will be reduced death in COVID-19 patient.

**Table 2.**
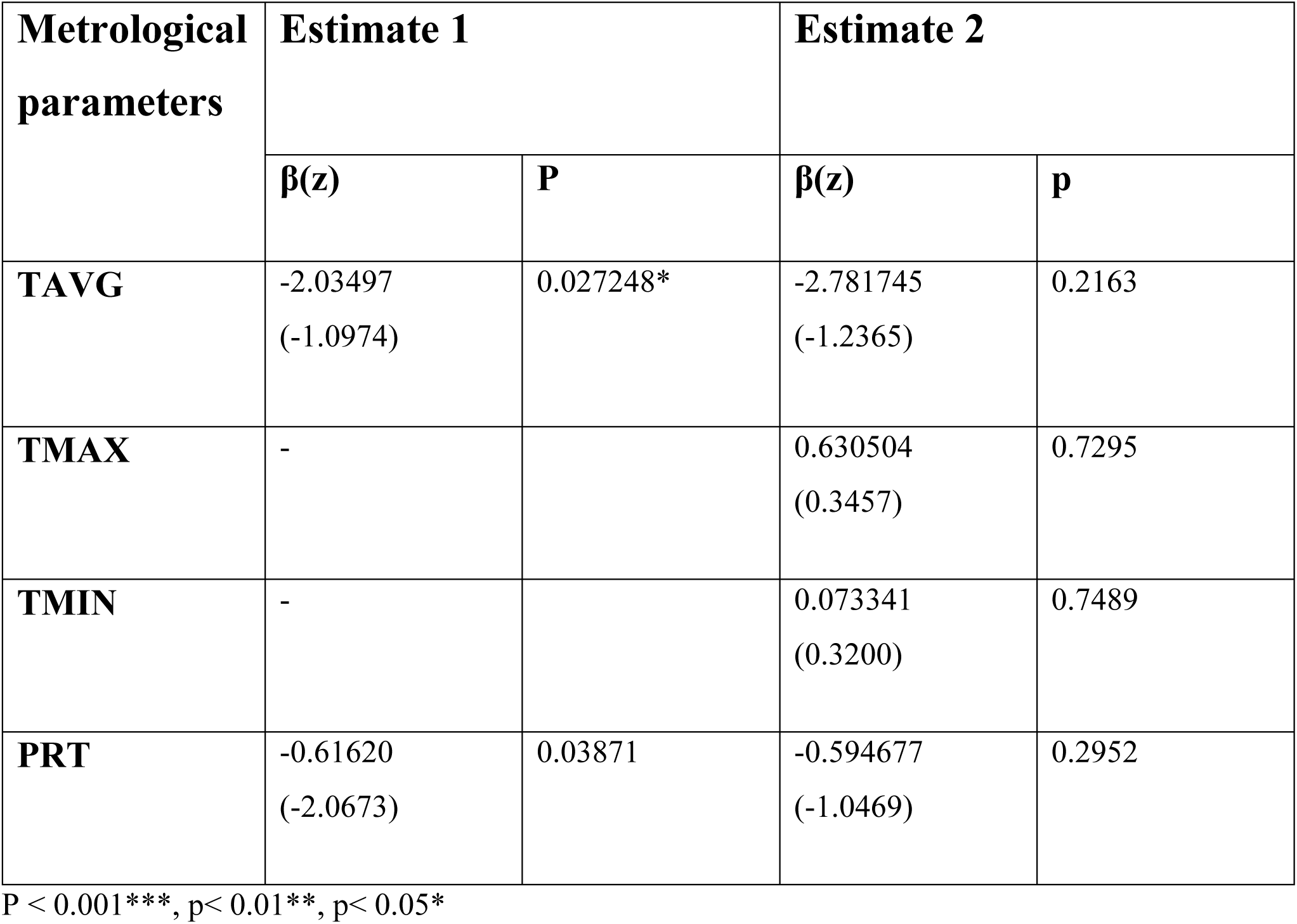
Outcome 1: Correlation of daily death and metrological parameter.

Table 3 shows the result of correlation analysis between daily new cases and metrological parameters. The first estimated model which includes average temperature and precipitation shows a negative correlation but the result doesn’t seem to be significant as the p value is more than 0.05. In the second estimated model with the inclusion of maximum temperature, minimum temperature along with average temperature and precipitation shows the positive correlation with average temperature and negative correlation with other parameters. The maximum temperature and average temperature seems to show the significant association with daily cases as the p value is less than 0.05. So the second estimated model shows as the temperature increases transmission of disease is reduced while transmission gets high during when temperature is reduced. The second estimated model also explains increase in the average temperature slightly increase the daily case.

**Table 3.** Outcome 2: Correlation of transmission and metrological parameters.

| **Metrological parameter** | **Estimate1** |  | **Estimate2** |  |
| --- | --- | --- | --- | --- |
|  | **$\beta$ (z)** | **p** | **$\beta$ (z)** | **p** |
| **TAVG** | -2.785<br>(-0.1234) | 0.901829 | 109.7364<br>(2.0227) | 0.04311* |
| **TMAX** |  |  | -103.5867<br>(-2.2724) | 0.02306* |
| **TMIN** |  |  | -1.8639<br>(-0.3205) | 0.74520 |
| **PRT** | -17.674<br>(-1.2521) | 0.210518 | -22.1401<br>(-1.5556) | 0.11980 |

P < 0.001\*\*\*, p< 0.01\*\*, p< 0.05

## Discussion

There are several studies which establish the fact that meteorological parameters do influence the respiratory virus, especially SARS-COV −2, which is the focus of interest in our study. We explored the association of meteorological parameters with daily new cases and daily new deaths, where our approach was focused on two estimate models. The first estimate model was used to find the association of daily new cases and daily new deaths with average temperature and precipitation, and in second estimate model, other two parameters (maximum temperature, minimum temperature) were also added with the former two parameters to find the association. For this we took daily new cases and daily new deaths of 7 populated cities across the world, when they were in the verge of community transmission or was already established having community transmission in order to rule out the incidence of high imported cases which could evidently impact our study of association.

We identified a significant association between temperature and daily new death due to SARS-COV −2. Average temperature was found to be inversely correlated with daily new deaths in the first estimated model. For every 1°c rise in average temperature there was a reduction in number of deaths by 2.03 deaths/day. This observation is similar with wu et al., who reported that there is a reduction in daily new deaths when the temperature rise. (Wu *et al*., 2020) Whereas, we observed that the average temperature was positively correlated with the transmission of disease and the association was significant. Even though this observation contradicts several studies like as that of Wang et al who observed a negative correlation with the rate of transmission and temperature,(Wang *et al*., 2020) there are studies such as of (Menebo 2020) which supports our findings. As per their hypothesis break ‘stay home’ rules by people when there is as a sunny climate could be one of the reasons of such a result. (Menebo, 2020) We found a potential relationship between maximum temperature and the rate of transmission. This is an impactful association, where we observed that for every 1°C rise in maximum temperature there is a reduction of number of cases by 103.58cases/day. Baker et al. had found a similar result, where they observed that the rate of transmission slows down as maximum temperature rises to around 50°F.(Sehra *et al*., 2020)This brings us to an inference that the materialization of warmer temperatures are likely to result in the fall of transmissibility of the virus. Our study substantiates the fact that temperature does have a role in decreasing the rate of transmission of COVID 19.

In case of precipitation we found a negative correlation with daily new deaths and no significant association with daily new cases. Baker et al., also reported that precipitation is not having any potential relationship with daily new cases.(Sehra *et al*., 2020) Smaller range or the decreased precipitation data obtained from the cities might have attributed to such an observation.

Due to the inaccessibility of proper data we have not taken humidity as a meteorological parameter in our study even, though certain studies proved humidity to have clear association with daily new cases and daily new deaths. Exclusion of other meteorological factors, Varied timing of lockdown rules, different health systems, improper reporting of daily news cases or daily new deaths in the cities and other environmental factors may have contributed to the limitation of our study.

Altogether, our study indicates that, there is a significant association of temperature with transmission of virus and death due to COVID-19.It is imperative to know the association which could be reflected upon local policy making, by considering other environmental as well as endogenous factors which could contribute to this association.

## Conclusion

This study investigates the effect of temperature (average temperature, maximum temperature and minimum temperature) and precipitation with daily mortality and transmission in the various cities around the globe which are been impacted most by the COVID-19. Our finding shows that increase in the average temperature partially reduces the daily death and increase in the maximum temperature helps to reduce the transmission. However, active measures must be taken to reduce the transmission of disease and control the source of infection.

## Data Availability

All the data have been collected from the public domain present in interner

## Acknowledgement

We are thankful for the support and help provided by our friends and teachers. No fund have been granted by the individual or the organization to support this study.

## Author Contribution

Amar Prashad Chaudhary: developed the topic, methodology and involved in manuscript drafting

Adna Nelson K : involved in manuscript drafting and data collection

Harish S: data analysis

S mythily: data collection and manuscript drafting

Chaithanya KJ: data collection and manuscript drafting

Lovely Nayak: data collection

Chiranjibi Sah: city selection

